# Pulmonary Hypertension: Intensification and Personalisation of Combination Rx (PHoenix): A phase IV randomised trial for the evaluation of dose-response and clinical efficacy of riociguat and selexipag using implanted technologies

**DOI:** 10.1101/2023.08.24.23294547

**Authors:** Frances Varian, Jennifer Dick, Christian Battersby, Stefan Roman, Jenna Ablott, Lisa Watson, Sarah Binmahfooz, Hamza Zafar, Gerry Colgan, John Cannon, Jay Suntharalingam, Jim Lordan, Luke Howard, Colm McCabe, John Wort, Laura Price, Colin Church, Neil Hamilton, Iain Armstrong, Abdul Hameed, Judith Hurdman, Charlie Elliot, Robin Condliffe, Martin Wilkins, Alastair Webb, David Adlam, Ray L Benza, Kazem Rahimi, Mohadeseh Shojaei-Shahrokhabadi, Nan X Lin, James M. S. Wason, Alasdair McIntosh, Alex McConnachie, Jennifer T Middleton, AA Roger Thompson, David G Kiely, Mark Toshner, Alexander Rothman

**Author notes:** **Corresponding author:** Alex Rothman, **Address:** IICD, The Medical School, Beech Hill Road, Sheffield, S10 2RX, United Kingdom, **Email:**.

## Abstract

Approved therapies for pulmonary arterial hypertension (PAH) mediate pulmonary vascular vasodilatation by targeting distinct biological pathways. Patients identified as intermediate-low risk, according to a four-strata risk assessment model, with an inadequate response to dual therapy with a phosphodiesterase type-5 inhibitor (PDE5i) and endothelin receptor antagonist (ERA), are recommended to either intensify oral therapy by adding a selective prostacyclin receptor (IP) agonist (selexipag), or switching from PDE5i to a soluble guanylate-cyclase stimulator (sGCS; riociguat). The clinical equipoise between these therapeutic choices provides opportunity for evaluation of individualised therapeutic effect. Traditionally, invasive/hospital-based investigations are required to comprehensively assess disease severity and demonstrate treatment benefit. Regulatory approved, minimally invasive monitors enable equivalent measurements to be obtained while patients are at home. In this 2x2 randomised crossover trial, patients with PAH established on guideline-recommended dual therapy and implanted with CardioMEMS™ (a wireless pulmonary artery sensor) and ConfirmRx™ (an insertable cardiac rhythm monitor), will receive ERA + sGCS, or PDEi + ERA + IP agonist. The study will evaluate clinical efficacy via established clinical investigations and remote monitoring technologies, with remote data relayed through regulatory approved online clinical portals. The primary aim will be establishing the change in right ventricular systolic volume measured by magnetic resonance imaging (MRI) from baseline to maximal tolerated dose with each therapy. Using data from MRI and other outcomes, including haemodynamics, physical activity, physiological measurements, quality of life, and side effect reporting, we will determine whether remote technology facilitates early evaluation of clinical efficacy, and investigate intra-patient efficacy of the two treatment approaches.

**Lay Summary:** This is a study to test if monitors placed in the lungs and the heart can help to choose the best medications for patients with a diagnosis of pulmonary arterial hypertension (PAH). PAH is a condition that results in high blood pressure in the blood vessels that supply the lungs. This study is for patients with PAH who are already taking two PAH medications (called dual therapy) but continue to have symptoms limiting their daily life and ability to exercise. There are two additional medications called selexipag and riociguat that may be prescribed when patients are not responding well to dual therapy; if selexipag is used, it is added to the existing dual therapy (Arm A), but if riociguat is used it replaces one of the drugs in the dual therapy (Arm B). It is not known which one of these treatment approaches is better. This 3-year study is called a crossover study design to look at responses of 40 individual patients to each of these two treatment approaches. Before starting the study treatment, patients will be implanted with monitoring devices. These approved devices will allow heart rate, pressure in the lungs, and other measures to be taken and seen by the clinical team from any location, without patients needing to attend hospital.

Patients will be randomly selected to either start Arm A or Arm B, as shown in **Figure 1**, before being swapped to the other treatment arm; there is no placebo. For example, for patients starting on Arm A, one of their medications (called phosphodiesterase type-5 inhibitor [PDE5i] e.g., sildenafil/tadalafil) will be stopped for a short time before riociguat is started. The dose of riociguat will gradually be increased to maximum dose and at 12 weeks they will have a magnetic resonance imaging (MRI) scan will measure treatment effect. The patient will then switch over to Arm B, where their PDE5i is restarted prior to treatment with selexipag. The MRI scan captures an image of the heart at the start and end of both Arm A and Arm B to look for improvements in the heart function with therapy. The study will also use patient-reported outcomes to record how patients feel and any side effects, blood tests related to heart health, and exercise tests to look at exercise ability. Each patient will be in the study for a total of 6 months, following which ongoing treatment choice will be decided at the discretion of the clinical care team.

---

Pulmonary arterial hypertension (PAH) represents a spectrum of disease that may be idiopathic or associated with genetic mutations, connective tissue disease, congenital heart disease, or drug/toxin exposure.^1^ At a cellular level, disease is driven by remodelling and constriction of the small pulmonary arteries, which can lead to right-sided heart failure and premature death.^2^ Currently available targeted therapies for this progressive disease mediate pulmonary vascular vasodilatation by acting on one of three pathways – the endothelin pathway via endothelin receptor antagonists (ERA), the nitric oxide (NO) pathway via phosphodiesterase type-5 inhibitors (PDE5i) or soluble guanylate cyclase stimulators (sGCS), or the prostacyclin pathways via prostacyclin analogues and prostacyclin receptor (IP) agonists.^1,3^ Despite the range of therapies available, drug choice is empirical and based on a hospital-based risk stratification that matches the number of vasodilator agents to disease severity.^3^

Approved therapies targeting the endothelin, NO or prostacyclin pathways have been shown to provide improvements in pulmonary vascular haemodynamics and 6-minute walk test (6MWT) in phase 2/3 studies.^1,4–8^ Evidence shows that time to clinical worsening is further improved in patients established on dual therapy with an ERA and a PDE5i, when compared with monotherapy with either agent.^3,9^ In line with European guidelines,^3^ for patients established on dual oral therapy (PDE5i/ERA) with an inadequate treatment response, NHS England’s National Commissioning Policy permits addition of the selective IP receptor agonist selexipag, or switching of PDE5i for the sGCS riociguat.^10,11^ There is clinical equipoise between these two approaches (i.e., triple therapy with selexipag + PDE5i + ERA and dual therapy with riociguat + ERA).

Selexipag, an IP receptor agonist, improves the time-to-clinical-worsening in patients on a range of background therapy regimens (including patients on no therapy [20.4%]; ERA or PDE5i monotherapy [47.1%]; and ERA/PDE5i dual therapy [32.5%]).^5^ However, data suggest that initiating triple therapy (PDE5i/ERA/IP receptor agonist) compared with dual therapy (PDE5i/ERA) in newly diagnosed treatment naïve patients offers no significant improvement in snapshot haemodynamic measurements or exercise capacity.^12^ Additionally, switching PDE5i for another drug targeting the NO pathway (riociguat, a sGCS) improves disease severity as assessed by World Health Organization (WHO) functional class and 6MWT distance, and reduces clinical worsening events compared with continuing PDE5i therapy.^2^ In all regulatory approval studies, significant side effects and therapeutic non-adherence were observed.^2,5,7,13–18^

In clinical practice, response to therapy and/or assessment of disease progression is made by assessing pressure and flow during invasive right heart catheterisation, by monitoring the downstream effect on right heart structure and function, and by evaluating exercise capacity. European Society of Cardiology (ESC) guidelines recommend invasive assessment of cardiopulmonary haemodynamics for disease diagnosis, assessment of severity, and to inform treatment decisions (therapeutic change and transplant).^3^ Other recommended means of guiding treatment decisions include regular measurement of exercise capacity by 6MWT, disease-specific risk scoring, assessment of right heart strain by N-terminal pro-brain natriuretic peptide (NT-proBNP), and/or non-invasive imaging, with magnetic resonance imaging (MRI) acknowledged to be more accurate than echocardiography.^3^

The current established standard for phase 2 studies of PAH therapies include assessment of invasive haemodynamics and the 6MWT. However, recent studies have demonstrated that non-invasive endpoints, such as right ventricular ejection fraction (RVEF) and right ventricular stroke volume (RVSV) measured by MRI, are repeatable, and detect treatment change in a manner similar to invasive catheterisation and NTpro-BNP, thereby establishing MRI as a robust, objective, non-invasive assessment of treatment response in patients with PAH.^19,20^

Despite these advances, there remains a need for invasive/hospital-based investigations to assess disease severity and demonstrate therapeutic benefit, and there are currently no means for early assessment of clinical efficacy in patients with PAH. This limits experimental medicine and drug development studies and prevents personalised medicine in clinical practice. Development of innovative approaches to monitor PAH outcomes is essential for a number of reasons including poor prognosis among patients with PAH, reduced quality of life, side effect profile of approved therapies, non-uniform drug response among patients, high cost of PAH-specific therapies (£5–120k/medication/patient/year), and emerging therapies with proven benefit in pre-clinical studies.^10,19–22^

In patients with PAH, cardiopulmonary haemodynamics are closely associated with clinical outcomes,^8^ and are affected by both disease worsening and increase or withdrawal of therapy.^23,24^ The development of minimally invasive technology that provides remote, daily measurement of cardiopulmonary haemodynamic parameters and physical activity may provide more comprehensive coverage of the effects of treatment on patients’ daily functioning, allowing insight into the intervening periods between scheduled hospital visits.^25^ Remote monitoring may provide benefit to both patients and care teams by allowing remote monitoring of efficacy following a treatment change. This may permit a personalised management approach, with the care team able to optimise therapy remotely – balancing therapeutic efficacy with side effects in each individual patient. Indeed, in patients with heart failure, remote, haemodynamic-guided therapy has been demonstrated to reduce heart failure hospitalisation,^26,27^ and these studies have led to regulatory approval of pulmonary artery pressure (PAP) monitors. Furthermore, studies of patients with PAH implanted with a PAP monitor and an insertable cardiac monitor (ICM) demonstrated that clinically indicated therapeutic changes altered physiological parameters associated with mortality, indicating that early, remote assessment of clinical efficacy may be achieved using these devices.^28^

CardioMEMS™ HF System (Abbott) is a wireless, CE-marked PAP monitor implanted at the time of right heart catheterisation to provide remote measurement of cardiopulmonary haemodynamics. CardioMEMS is approved for routine clinical practice in the USA and Europe, and to date over 30,000 of these monitors have been implanted. The Confirm Rx™ ICM (Abbott) is a minimally invasive CE-marked, FDA-approved cardiac monitor, implanted in a clinic setting for patients who experience unexplained symptoms, such as dizziness; palpitations, chest pain, syncope, shortness of breath, as well as for patients who are at risk for cardiac arrhythmias. Over 40,000 Confirm RX™ have been implanted and the device is in routine clinical use in the UK.

Here, we detail the protocol for a study in which patients with intermediate-low risk PAH, established on guideline-recommended dual oral therapy,^29^ will be implanted with CardioMEMS and ICM devices. Following implantation, patients will enter a 2x2 crossover study in which PDEi will be replaced with sGCS (ERA + sGCS), or an IP receptor agonist will be added to PDEi and ERA (PDEi + ERA + IP receptor agonist). Data obtained from remote monitoring will be compared with that from established clinical investigations undertaken at baseline and maximal therapy on each drug. The crossover design of this study, which will incorporate structured up-titration of these drugs, is aimed at evaluating the capacity of implantable technology for early evaluation of the clinical efficacy of these drugs. A crossover study is the logical study design to investigate intra-patient efficacy of these treatment options and increases the power to detect clinical efficacy. Additionally, the study will provide insight regarding the capacity of remote monitoring technology to facilitate trials that are not currently possible due to the requirement for repeated, hospital-based invasive/imaging procedures.

## Methods

### Study design

This open-label, phase 4, multicentre, randomised 2x2 crossover study (NCT05825417) in patients with PAH established on dual therapy (PDE5i/ ERA) will evaluate the effects via clinical investigations, patient-reported outcomes and remote cardiac monitoring of two therapeutic strategies – adding an oral drug targeting the prostacyclin pathway (selexipag; PDEi + ERA + IP receptor agonist) and switching of PDE5i to an sGCS (riociguat; ERA + sGCS). Using the 2x2 crossover trial design, patients will receive both therapies but the sequence will be randomly assigned with washout phases between therapies, and assessments of response to each therapy to be performed.^30^ The study protocol was approved by the NHS Health Research Authority (IRAS PROJECT ID 325120, REC Reference 23/NE/0067). A tabulated summary of all visits and assessments is provided in **Supplementary Table 1**.

### Objectives and endpoints

The main aims of this study will be to assess the individual difference in effect between treatment escalation with selexipag (PDEi + ERA + IP receptor agonist) or riociguat (riociguat; ERA + sGCS) on RV stroke volume (flow) as measured by cardiac MRI in patients with intermediate-low risk PAH, and to determine whether remote monitoring devices can be used to provide an early assessment of clinical efficacy of drug therapies for PAH.

The primary endpoint will be change in RVSV (flow) measured by MRI from baseline to 12 weeks for each therapeutic strategy. Change in RVSV provides a robust, objective assessment of clinical efficacy and will represent a clinically meaningful change in physiology.^31^

Secondary endpoints for each therapeutic strategy include change from baseline to 12 weeks in haemodynamics (total peripheral resistance [TPR], mean pulmonary artery pressure [mPAP], cardiac output [CO], cardiac index, stroke volume [SV], heart rate [HR]), 6MWT, NTpro-BNP, MRI parameters (RVEF, right ventricular end systolic volume [RVESV], right ventricular end diastolic volume [RVEDV], RVSV (volume), left ventricular ejection fraction [LVEF], left ventricular end systolic volume [LVESV], left ventricular end diastolic volume [LVEDV], LVSV flow), quality of life (EmPHasis-10), medication compliance (PHoenix PRO) and side effects, depression and anxiety symptoms (Generalized Anxiety Disorder-[GAD]-2/7 and Patient Health Questionnaire [PHQ]-2/9), WHO functional class, and activity levels as measured with a Garmin Venu2 smartwatch. A full list of outcome measures is provided in **Table 1**.

**Table 1.** Outcome measures.

|  | Hospital-based measures | Remotely monitored measures |
| --- | --- | --- |
| Primary outcomes | Change in RVSV (flow) measured by MRI |  |
| Secondary outcomes | Haemodynamics:<br>• TPR<br>• mPAP<br>• CO<br>• Cardiac index<br>• SV<br>• HR<br>MRI parameters:<br>• RVEF<br>• RVESV<br>• RVEDV<br>• RVSV (volume),<br>• LVEF<br>• LVESV<br>• LVEDV<br>• LVSV flow<br>Other parameters: | Confirm Rx:<br>• HR<br>• HR variability<br>• Cardiac rhythm<br>• Thoracic impedance<br>• Respiratory rate<br>CardioMEMS:<br>• mPAP<br>• CO<br>• SV<br>Smartwatch (Garmin Venu2):<br>• Physical activity<br>• Physiological measurements (TPR, mPAP, CO, cardiac index, SV, and HR) |
|  | • 6MWT<br>• NTpro-BNP<br>• WHO functional class<br>• Quality of life (EmPHasis-10)<br>• Depression and anxiety symptoms (GAD-2/7 and PHQ-2/9)<br>• Medication compliance (PHoenix PRO questionnaire)<br>• Side effects | Phone apps:<br>• PRO (Phoenix remote questionnaire)<br>• Medication compliance (myCardioMems app)<br>6MWT |

6MWT, 6-minute walk test; CO, cardiac output; HR, heart rate; GAD, Generalized Anxiety Disorder; LVEF, left ventricular ejection fraction; LVEDV, left ventricular end diastolic volume; LVESV, left ventricular end systolic volume; mPAP, mean pulmonary artery pressure; PHQ, Patient Health Questionnaire; PRO, patient-reported outcome; RVEDV, right ventricular end diastolic volume; RVESV, right ventricular end systolic volume; RVSV, right ventricular stroke volume; SV, stroke volume; TPR, total peripheral resistance.

Established clinical study endpoint measures at maximal therapy will be compared to changes in remotely monitored parameters measured at 4 and 8 weeks to determine if the implanted devices can detect structured changes in the clinical therapy, thereby facilitating early assessment of clinical efficacy. Remote monitored parameters to be correlated with maximal therapy assessments, measured as absolute change from baseline and area under the curve to 4 and 8 weeks on each therapy, include haemodynamics (mPAP, CO, cardiac index, TPR, day HR, night HR, HR reserve), activity (minutes per day), 6MWT, and PRO.

Analysis will also be performed to determine if changes in established and remotely monitored parameters (primary and secondary endpoints) can detect individual patient-level therapy effects, thereby determining the utility of remote monitoring for personalised treatment plans.

### Study population

The study aims to recruit 40 patients with PAH, established on PDE5i and ERA, through the UK National Pulmonary Hypertension Clinical Studies Network (UNIPHY) – a collaboration of UK centres commissioned for the treatment of PAH, providing access to all patients within the UK currently receiving targeted therapy for PAH (>5000).^33,34^ Suitable patients will be identified from existing patient lists by local teams and invited to screening via clinical and research teams.

Eligible patients meeting the inclusion and exclusion criteria (**Table 2**) will be established on PDE5i/ERA dual therapy and will meet NHS England’s National Commissioning and ESC guideline criteria for initiation of IP receptor agonist or sGCS.^3,10,11^ While it is expected the majority of patients recruited will have intermediate-low risk PAH, patients with intermediate-high risk PAH who decline intravenous therapy will also be considered.

**Table 2.**
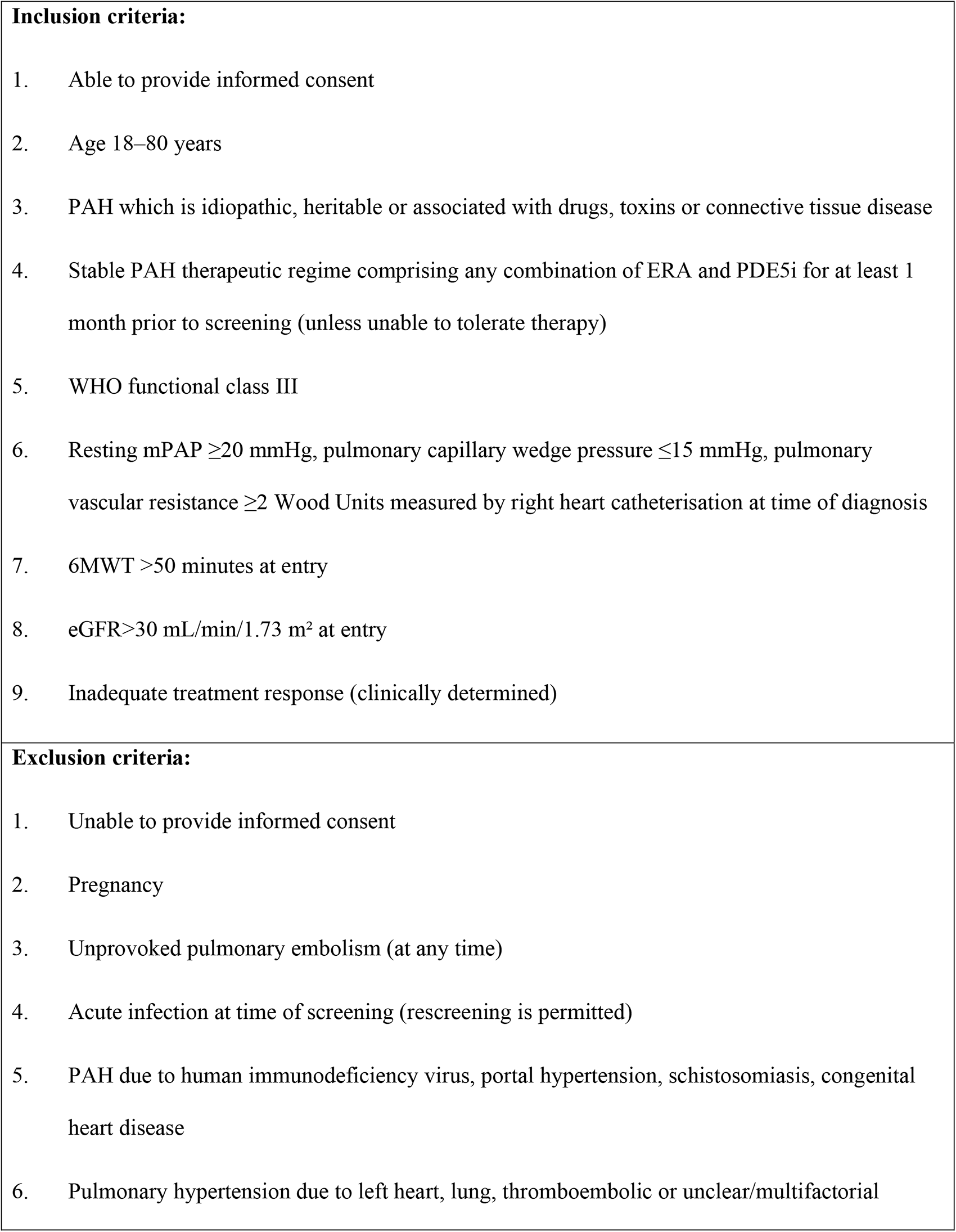
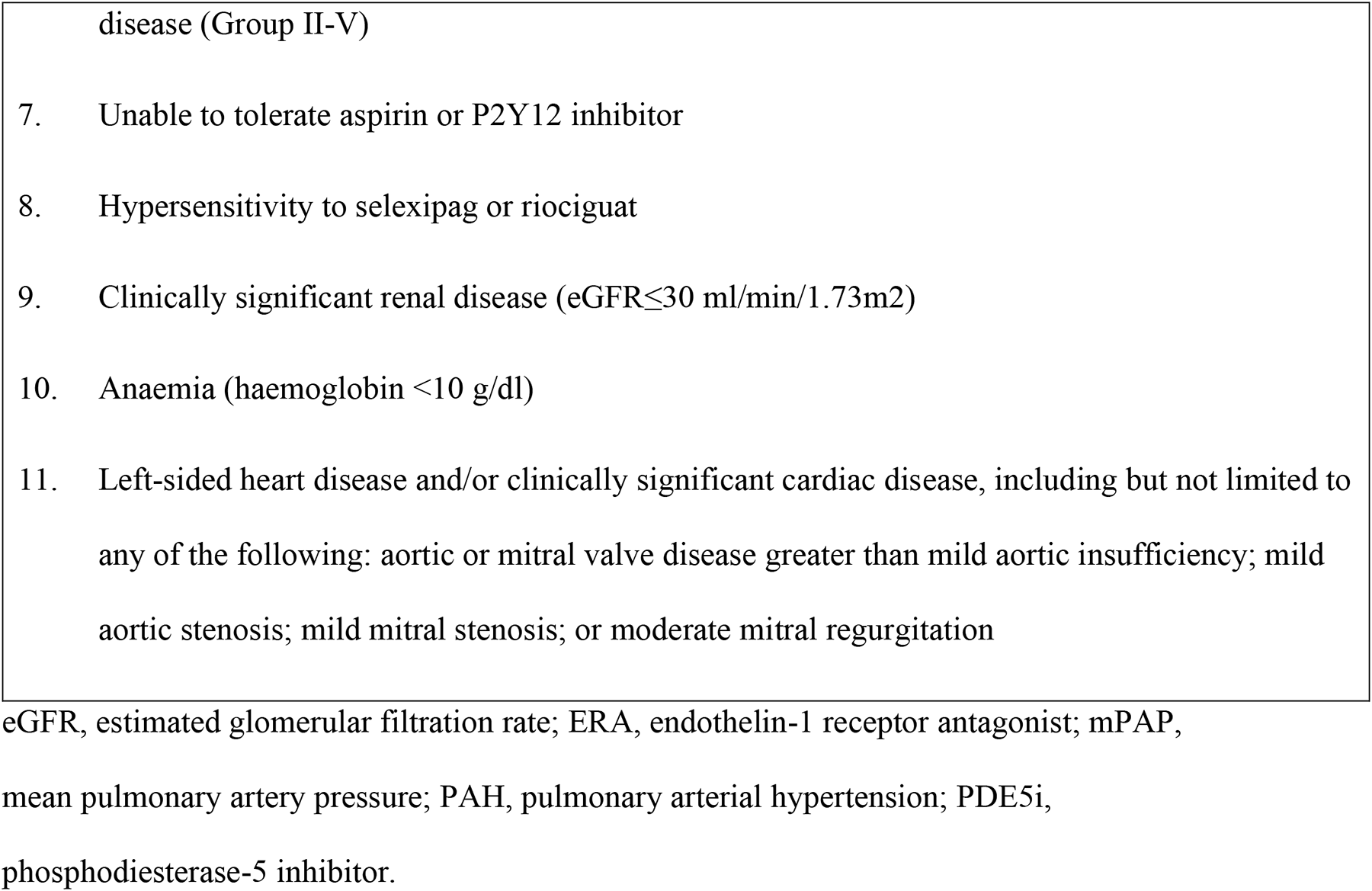
Summary of eligibility criteria.

### Device implantation

Eligible patients will be implanted using standard techniques with CardioMEMS and Confirm Rx ICM devices and remote monitoring data collected using regulatory-approved online portals.^28^

### Treatment

Patients will be randomised 1:1 to one of two treatment sequences (**Figure 1**), with comparisons to be made using patient-level data within the two treatment arms. Randomisation will be done by authorised staff at study sites using a concealed randomisation system via the Zeesta electronic case report form (eCRF – www.zeesta.ai/) portal. A block randomisation stratified by site with a block size of four will be employed.

As per standard practice, the treatment schedules will include a minimum PDE5i washout period of 24–48 hours (depending on PDE5i) prior to initiation of riociguat;^2^ in addition, riociguat and selexipag will be titrated according to established dose-adjustment schemes over a 6-week period to maximum doses of 2.5 mg three times per day, and 1600 μg twice daily, respectively as tolerated.

In brief, patients in Arm A will initiate treatment with selexipag for uptitration to maximal therapy. Prior to crossover, patients will undergo selexipag dose de-escalation and washout, followed by PDE5i washout. Patients will then initiate treatment with riociguat for uptitration to maximal therapy. Patients in Arm B will have an initial PDE5i washout period before commencing treatment with riociguat. This will be followed by de-escalation and washout, and initiation of treatment with PDE5i prior to crossover to selexipag treatment. In both arms the primary endpoint evaluation will be undertaken following a minimum intended duration of 5 weeks on maximal tolerated dose of each therapy. The dose escalation and de-escalation protocol are shown in **Supplementary Table 2**.

### Clinical assessments

Patients will undergo clinical assessments at baseline before receiving study drug treatment, and at Week 12 and Week 27 of the treatment schedule (**Figure 1**); these will include haemodynamics, 6MWT, MRI and NT-proBNP assessments.

MRI analysis will be provided by a study-appointed core lab using certified clinicians; scans will be de-identified and analysed in random order independent of patient and time point. Analysis of primary and secondary endpoints will be undertaken in a blinded manner by an independent statistical team in accordance with a pre-specified statistical analysis plan.

### Remote monitoring

Physiological parameters to be monitored will include TPR, mPAP, CO, SV, and HR (**Table 1**). Patients will be given instructions on how to take readings at the time of implantation. In addition, remote detection of changes in physical activity levels will be measured by Garmin Venu2 smartwatch, and the 6-minute walk test (6MWT) performed at home.

### Patient-reported outcomes

To date, no published randomised controlled trials in PAH have undertaken PAH-specific PRO instruments as secondary endpoints (**Figure 2**). The current study aims to understand patients’ attitudes about PAH medications and the impact of the study medication on quality of life, as well as exploring attitudes about the use of remote technologies.

**Figure 1.**
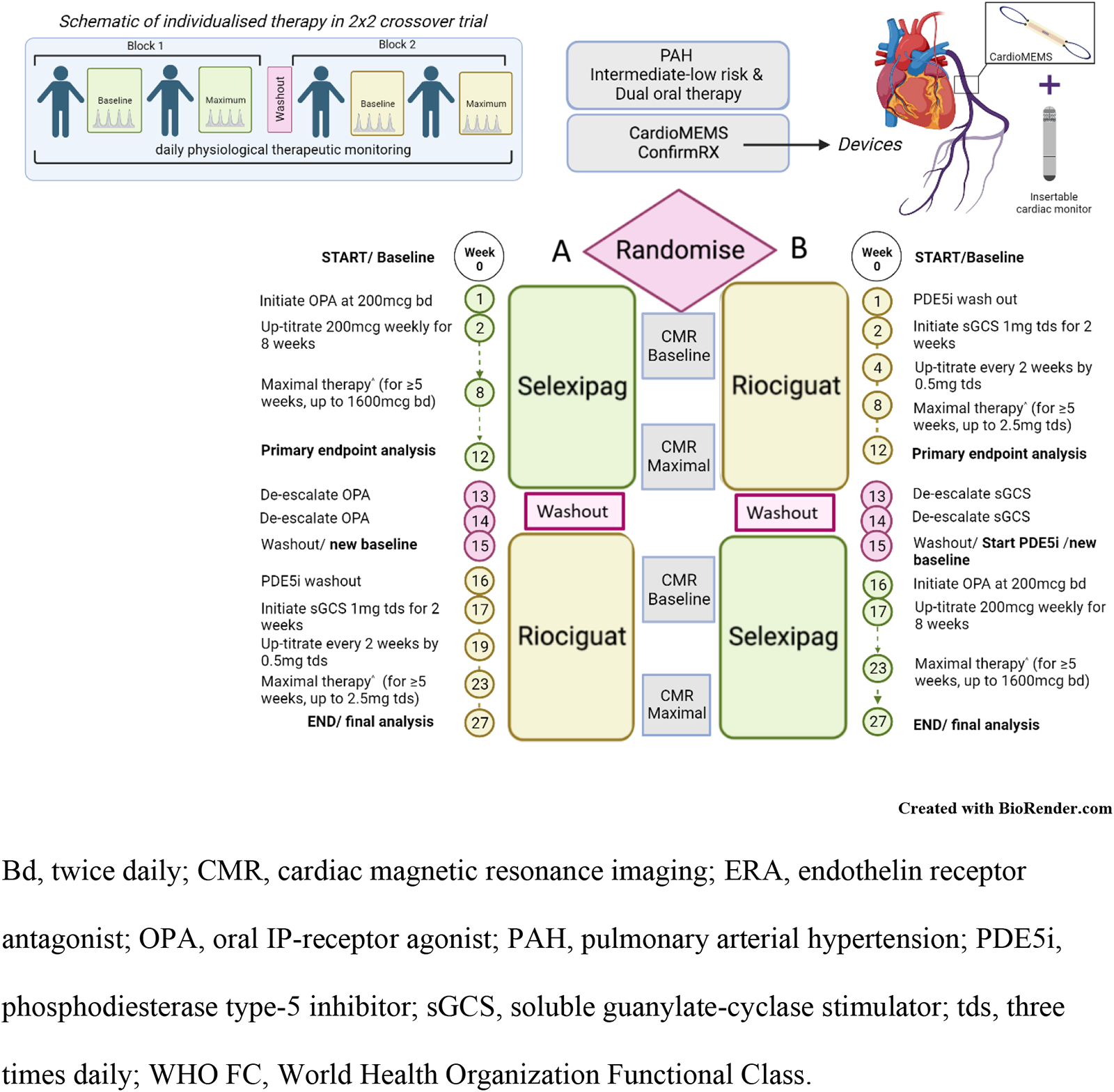
Dose escalation and de-escalation protocol

**Figure 2.**
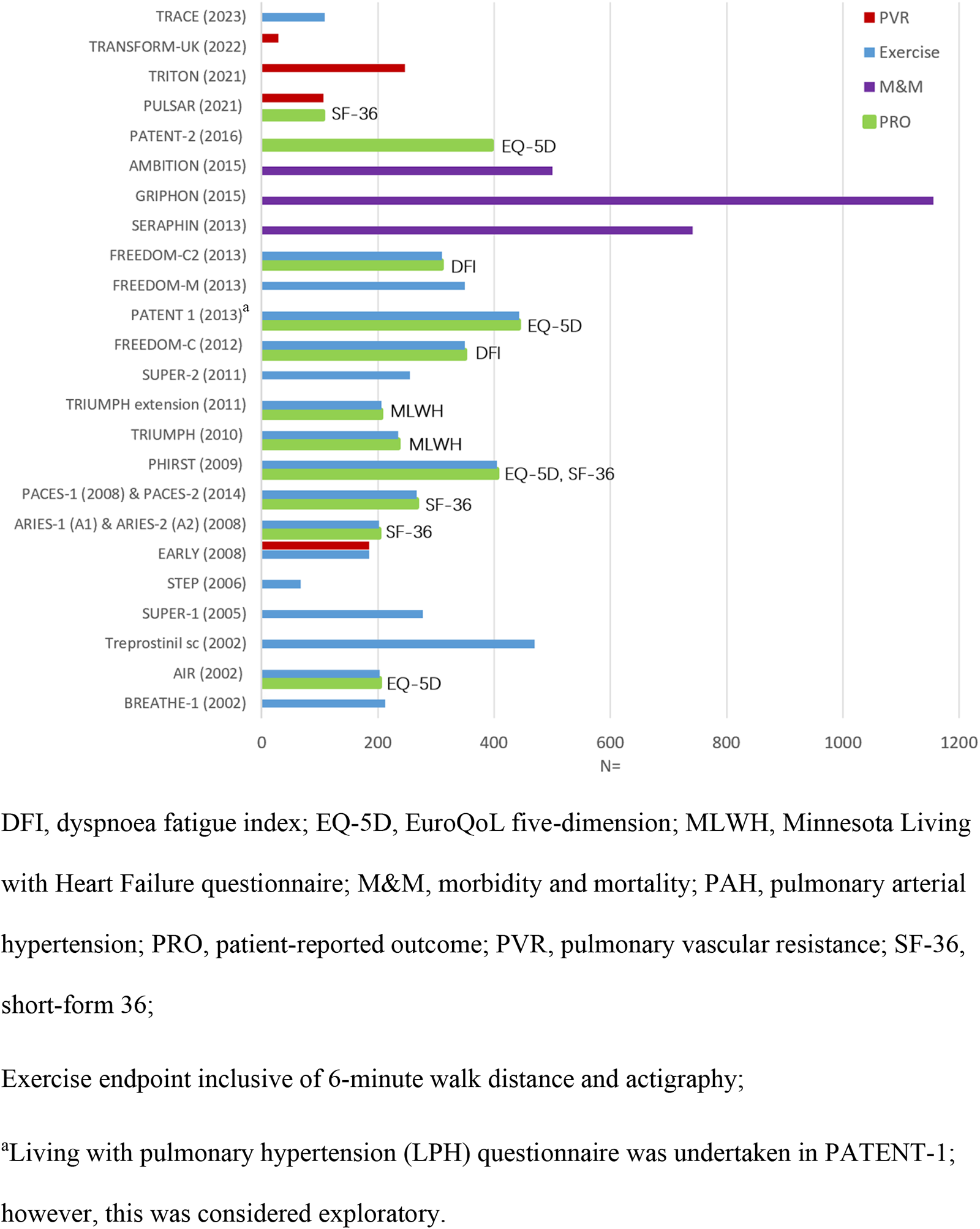
Primary outcomes in landmark PAH randomised controlled trials with patient-reported outcomes included as secondary endpoints

Quality of life outcomes will be assessed weekly using the validated EmPHasis-10 questionnaire.^35^ Validated questionnaires will be used twice monthly to screen for anxiety (GAD-2)^36^ and depression (PHQ-2) symptoms.^37^

Patients will also be asked to record, on a weekly basis, any side effects of the study medications and to track dose-response changes that are observed. Data will be collected on patients’ attitudes towards their PAH medications and patient-reported medication compliance (PHoenix PRO questionnaire; **Supplementary** Figure 1). All participants will be invited to co-enrol in the COHORT-digital study,^38^ which enables PRO reporting through a mobile application called Atom5™ (**Figure 3**). If participants decline to co-enrol for digital PRO reporting, data for these outcomes will be collected using a 10-item questionnaire via telephone communication into the eCRF.

**Figure 3.**
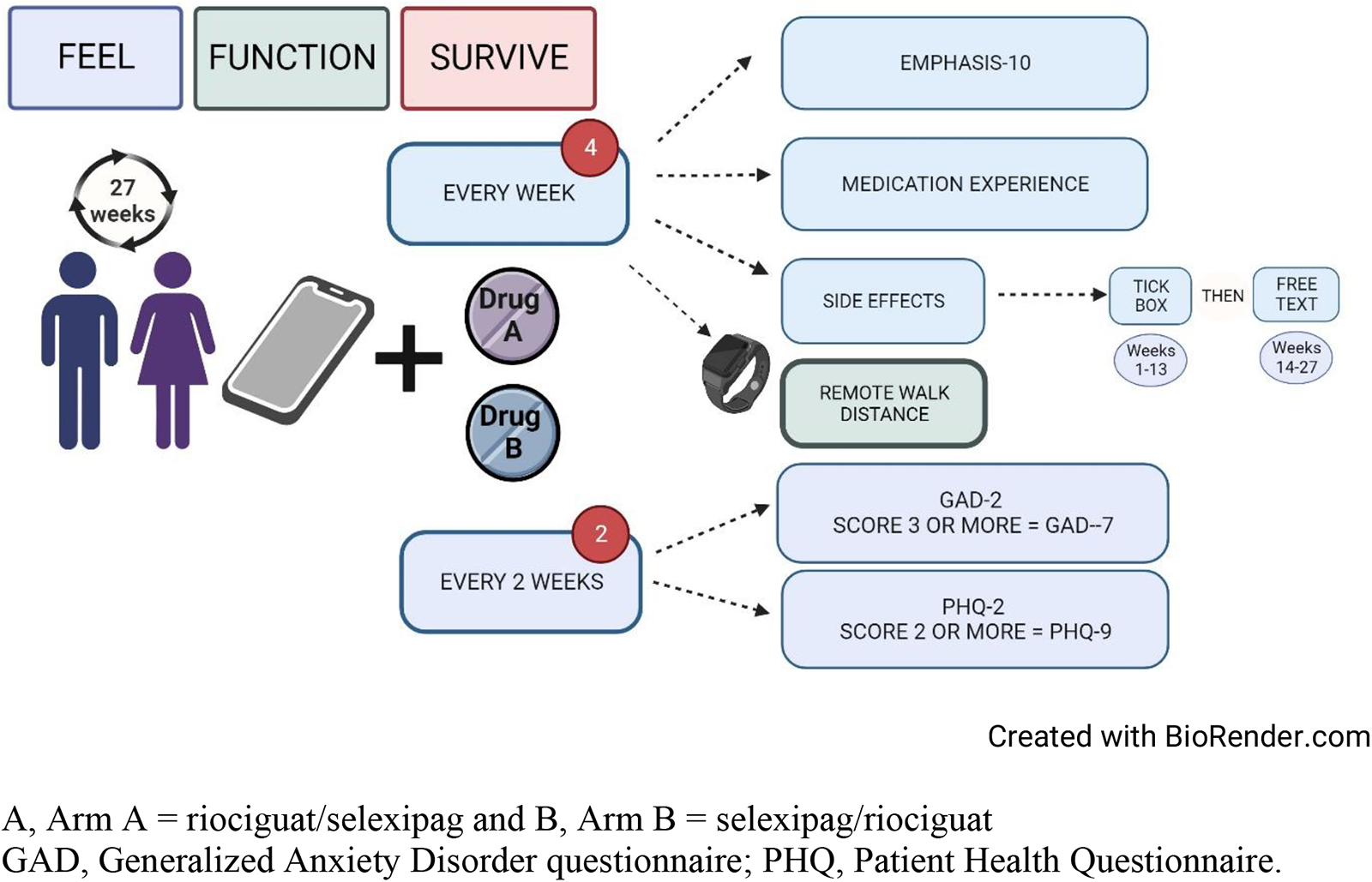
Patient-reported outcomes in the PHoenix study

Additionally, patients will also be asked to provide their insight to help understand attitudes regarding remote monitoring and clinical care and health outcomes at enrolment and study completion.

### Safety

Adverse events (AEs) will be monitored over the duration of the study period; AEs that are definitely or possibly related to the device or the insertion procedure should be considered device-related (adverse device effect [ADE]).

### Statistical analyses

#### Sample size calculation

A sample size was chosen to ensure adequate power to detect differences in the clinical efficacy of the two treatment approaches in population-level analyses and to have adequate power to evaluate the ability of implantable/remote technology to provide early evaluation of such clinical efficacy. For comparing the clinical efficacy of the two treatment approaches, we have used published RVSV data reporting a minimal clinically important difference of 12 mL and within-patient standard deviation (SD) of 16.5 mL.^39^ The current study will be powered using a standardised effect (SE) of 12/16.5=0.73. Assuming 1:1 randomisation of the participants to the two treatment sequences, 40 participants will provide 90% power in the population-level clinical efficacy analysis for a 5% two-sided type-I error-rate, with SE of 0.73. This is below the SE previously reported for the pulmonary vascular resistance (PVR; SE=353.4/219.0=1.61),^4^ and the RVEF (SE=9.12/7.39=1.23)^20^ and comparable to that observed for the 6MWT (SE=36.0/46.7=0.77).^7^ Therefore, the study will be well powered for population-level analyses of these additional important outcomes.

The sample size of 40 also provides good power for assessing whether changes in remote physiological measures from baseline to 4 and 8 weeks (mPAP, CO, HR, and heart rhythm) are correlated with change in clinical measures from baseline to 12 weeks. The sample size of 40 patients provides 90% power, at a two-sided 5% type-I error-rate, for correlations greater than 0.5, which would represent those of clinical interest.

The study is not powered for formal mediation analysis, so this will be considered exploratory. No formal multiple testing correction will be applied.

#### Statistical analysis plan

All statistical analysis plans (SAPs) will be drafted early in the study and finalised prior to the analysis of unblinded data.

The primary clinical efficacy analysis will use a linear mixed effects model with the dependent variable being the change in RVSV (flow) from baseline to 12 weeks. “Baseline” and “12 weeks” refer to the time points within each treatment period. Each participant will contribute up to two observations if they complete both treatment periods. A random effect for each participant will be included together with the within-treatment period baseline RVSV (flow) measurement, a fixed period effect, and treatment (selexipag or riociguat) allocated during the treatment period. This model will be used to estimate the mean difference between the two treatment approaches (ERA + sGCS and PDEi + ERA + IP receptor agonist), together with 95% confidence interval and a p-value using a Wald test.

Similar methods will be applied to the analysis of secondary efficacy outcomes. Treatment effect heterogeneity between subgroups will be assessed by including treatment-by-subgroup interaction terms in regression models. Main analyses will use complete data only, but multiple imputation will be used for missing data in sensitivity analyses. No adjustments will be made for multiple statistical testing. All efficacy analyses will follow the intention to treat principle (i.e., analysis according to randomised treatment, regardless of treatment compliance). Safety data (adverse events and side effects) will be summarised in relation to treatment being received at the onset of the event, and the study period (pre-treatment period 1, treatment period 1, washout period, treatment period 2, post-treatment period 2); no formal statistical comparisons will be applied.

To analyse whether changes in RVSV (flow) may be explained by remotely monitored physiological parameters, we will test whether there is a significant correlation between change between baseline and Week 4/Week 8 physiological parameters and change between baseline and Week 12 RVSV (flow) outcome. We will also adopt a mediation analysis approach to investigate what proportion of the change between baseline and Week 12 RVSV (flow) is explained by the change between baseline and Week 4/Week 8 physiological parameters using the mediation package in R. Secondary endpoints will be analysed using appropriate regression models.

## Discussion

Patients with PAH receiving dual combination treatment (PDE5i + ERA) who are stratified as being at intermediate-low risk are recommended to intensify therapy via addition of the IP receptor agonist, selexipag, or to switch from a PDE5i to the sGCS, riociguat.^10,11,29^ There is clinical equipoise between triple therapy with selexipag + PDE5i + ERA and dual therapy with riociguat + ERA. Conducting head-to-head clinical trials to compare treatment strategies in patients with PAH is challenging due to the current necessity for repeated, hospital-based invasive/imaging procedures to evaluate treatment efficacy. Remote haemodynamic and cardiac monitoring may provide a means for early, minimally invasive evaluation of clinical efficacy and early identification of clinical worsening in patients with PAH, which may facilitate study designs evaluating dose-response, time-to-effect and head-to-head comparison. This study is designed to assess the individualised effect of selexipag and riociguat on RV stroke volume as measured by cardiac MRI, and to determine whether remote monitoring devices can be used to provide an early assessment of clinical efficacy of drug therapies for PAH. This hybrid drug-device regulatory approved design represents the first evaluation of an sGCS and IP agonist.

This blinded analysis of MRI objective measures provides an efficient, robust and effective clinical study structure. As the primary endpoint is objective with blinded analysis, patients and clinicians will not be blinded to the sequence of drug allocation and up-titration.

In this study, data will be relayed daily from regulatory approved, minimally invasive monitors to care teams through secure online clinical portals, with the aim of facilitating early, individual-level, remote evaluation of treatment effects. This study will offer the potential to build on existing evidence showing that remotely monitored parameters may be used at diagnosis to categorise patients with PAH as low, intermediate, or high risk.^24^ In addition to offering the potential for early evaluation of therapies, the use of remotely monitored outcomes may provide a broader picture of the effects of treatment on patients’ daily functioning.^25^ Remote patient monitoring may also facilitate more patient-centric research, and improve study recruitment and retention, which are key issues for research into a rare disease such as PAH.^21,25^ Additionally, patients with PAH are typically prescribed combination therapies, which can make it challenging to power trials to demonstrate the effectiveness of novel therapies.^20,21^

In addition to capturing data on established physiological and biochemical markers of clinical risk, this study will provide valuable insight into patient-reported quality of life and mental health outcomes, as well as exploring side effects experienced during therapeutic up-titration and withdrawal of therapy.

## Data Availability

Methods paper therefore not applicable.

## Declarations

Dr Frances Varian: MRC clinical research fellow, conference funding from Janssen and Janssen

Dr Jennifer Dick: none declared

Mr Christian Battersby: none declared

Mr Stefan Roman: none declared

Miss Jenna Ablott: none declared

Dr Lisa Watson: none declared

Mrs Sarah Bizmahfooz: none declared

Dr Hamza Zafar: none declared

Dr Gerry Colgan: no direct conflicts, has undertaken consultancy work & honoraria for Janssen Ltd, Bayer Ltd, MSD. Received research funding from Janssen Ltd.

Dr John Cannon: support to attend conferences from Janssen and been paid for advisory boards by Janssen and Ferrer.

Jay Suntharalingam: none declared

Dr Jim Lordan: none declared

Professor Luke Howard: I have received honoraria for advisory boards, steering committees and speaking from Janssen. My department has received research funding support from Janssen. I have received personal support for travel, accommodation and registration at international meetings. I have received honoraria for advisory boards and speaking from MSD. I have received honoraria for advisory boards from Endotronix.

Colm McCabe: none declared

Dr John Wort: I have received honoraria from Janssen, MSD, Ferrer and Acceleron, research grants from Janssen and Ferrer and travel and accommodation grants from Janssen

Laura Price: none declared

Dr Colin Church: none declared

Dr Neil Hamilton: Honoraria from MSD and Janssen, travel and accommodation grants from Janssen, participation on advisory boards for Bayer, MSD, Janssen and Vifor and is a board member on the NHS Specialist respiratory clinical reference group. Iain Armstrong

Dr Abdul Hameed: none declared, https://orcid.org/0000-0002-2128-2081

Dr Judith Hurdman: none declared

Dr Charlie Elliot: none declared

Prof Robin Condliffe: No COI. Received honoraria for speaking and advisory boards from Janssen and MS

Prof Martin Wilkins: support from NIHR for clinical research facility and biomedical research centre infrastructure support BHF centre support (RE/18/4/34215), consulting fees for MorphogenIX, Janssen and Janssen, Kinaset, Chiesi, Aerami, BenevolentAI, Novartis, and VIVS, participation on data safety monitoring board for Acceleron and GSK, Associate Professor Alastair Webb: none declared

Dr David Adlam: none declared

Professor Ray L Benza: steering and adjudication committees ABBOTT

Professor Kazem Rahimi: receives grants from the Oxford Martin School and the British Heart Foundation. KR is an associate editor of Heart and specialty editor of PLOS Medicine. KR is a co-founder of Zeesta and sits on the advisory board of Medtronic

Dr Moha Shojaei: none declared

Dr Nan Lin: none declared

Prof James Wason: none declared

Dr Alasdair McIntosh: none declared

Prof Alex McConnachie: none declared

Dr Jennifer Middleton: none declared

Dr Roger Thompson: I have received honoraria, travel support and grant funding from Janssen Prof David Kiely: Support and grants received from NIHR Sheffield Biomedical Research Centre, Janssen Pharmaceuticals; additional grants from Ferrer; consulting fees, honoraria payments and supports for attending meetings received from Janssen Pharmaceuticals, Ferrer, Altavant, MSD and united Therapeutics, participants on advisory boards with Janssen and MSD; members of clinical reference group for specialist respiratory medicine (NHS England) and lead of UK national audit of pulmonary hypertension.

Dr Mark Toshner: Funding from NIHR Cambridge BRC, NIHR HTA; consulting fees from MorphogenIX and Jansen, participation on data safety monitoring board/advisory board with ComCov and FluCov.

Dr Alex Rothman: Research funding: Wellcome Trust Clinical Research Career Development Fellowship (206632/Z/17/Z), Medical Research Council (UK) Experimental Medicine Award (MR/W026279/1), NIHR Biomedical Research Center Sheffield, Contribution in kind: Medtronic, Abbott, Endotronix, Novartis, Janssen. Research support and consulting: NXT Biomedical, Endotronix, SoniVie, Neptune, Gradient. MT: Research funding: NIHR Biomedical Research Center Cambridge, NIHR HTA. Personal support: GCK and Jansen. DSMB: ComCov, FluCov. PDM: personal support: Abbott.

## Ethical approval

IRAS project ID: 325120

REC reference: 23/NE/0067

## Contributorship

The study was designed by AR with representatives from the UK Pulmonary Hypertension Network (CC, GC, RC, LH, DK, JL, AR, JS, MT and SJW), the clinical research fellow (FV) and the clinical trial manager (JD). All the authors contributed to the development of the manuscript and approved the final version of the article.

## Acknowledgements

The authors thank Aisling O’Keeffe, PhD, and Allison Kirsop, PhD, of Scientific Writers Ltd., for providing medical writing support.

## Supplementary appendix

**Supplementary Table 1.**
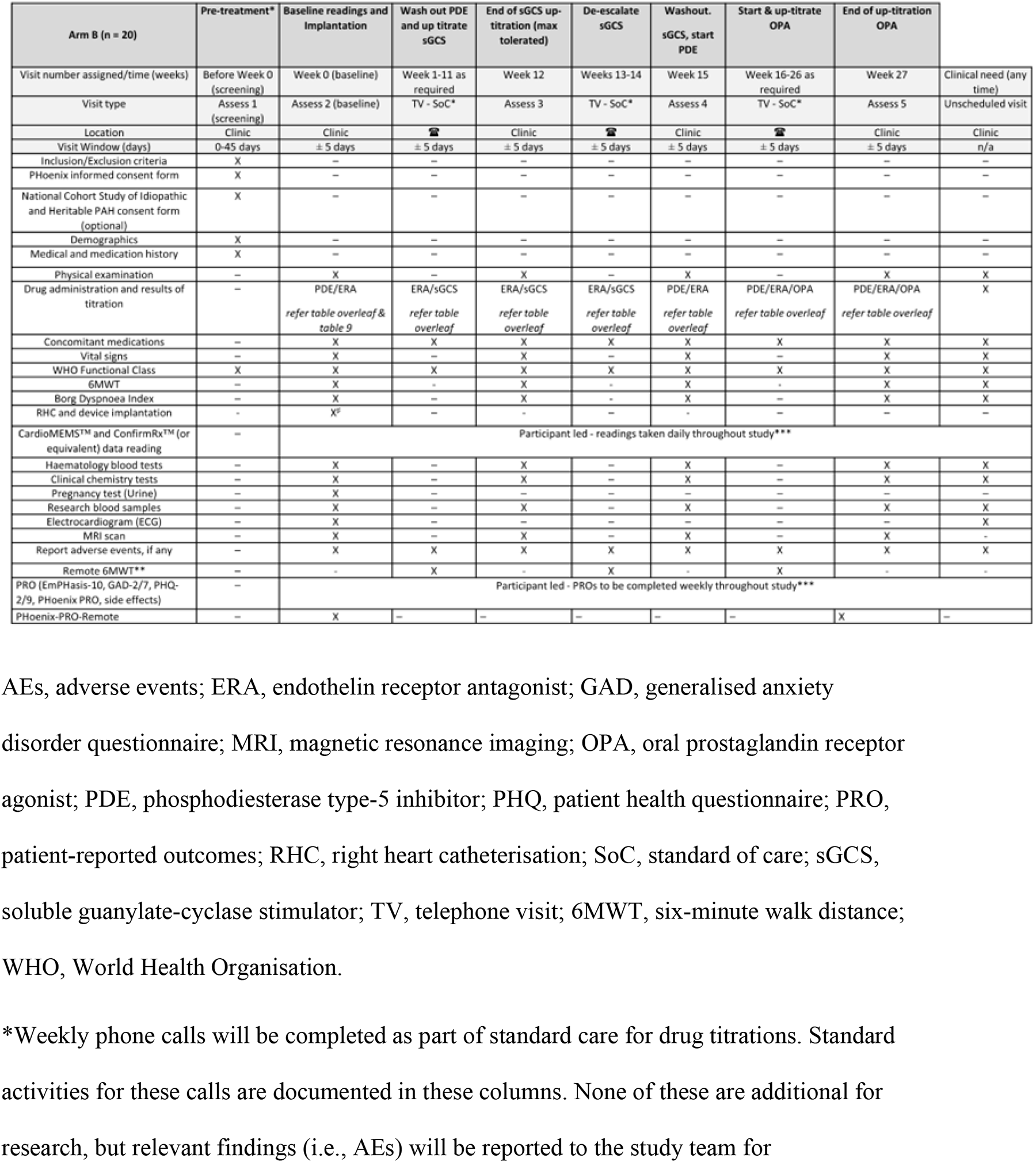
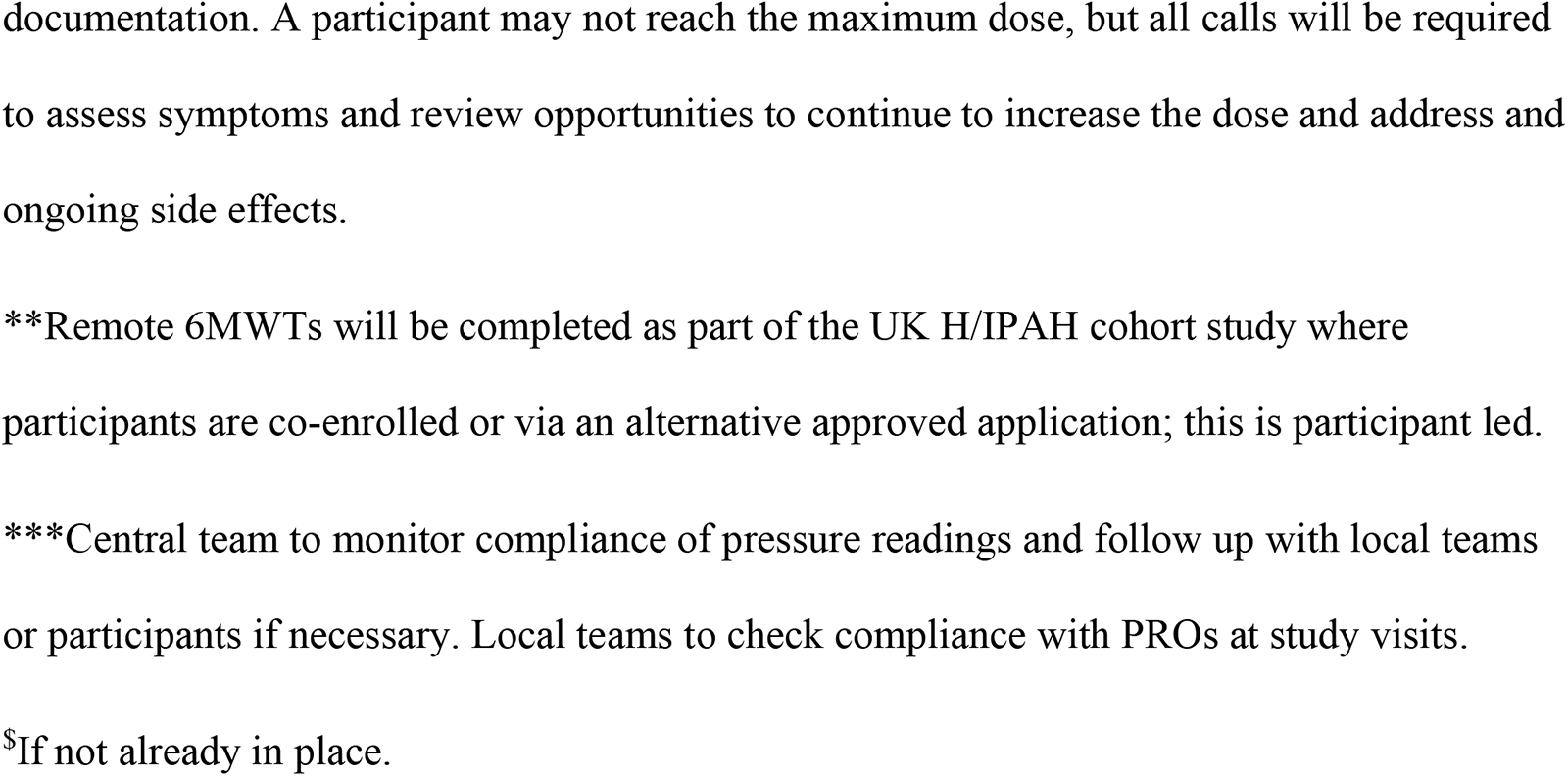
Schedule of events Assessments at screening and baseline need only be completed once if no change in clinical condition between visits. Dosing schedule follows on next page.

**Supplementary Table 2.**
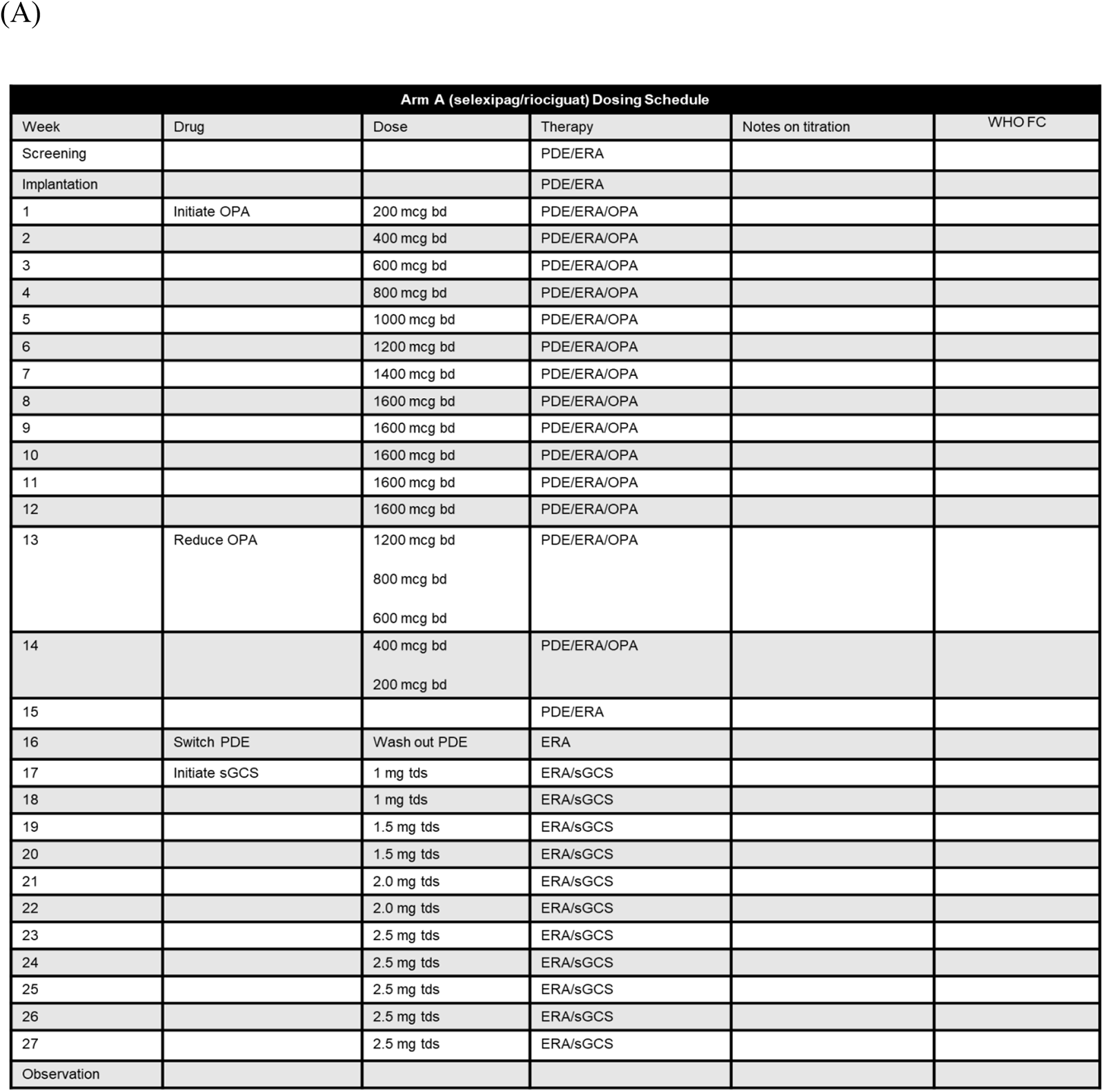
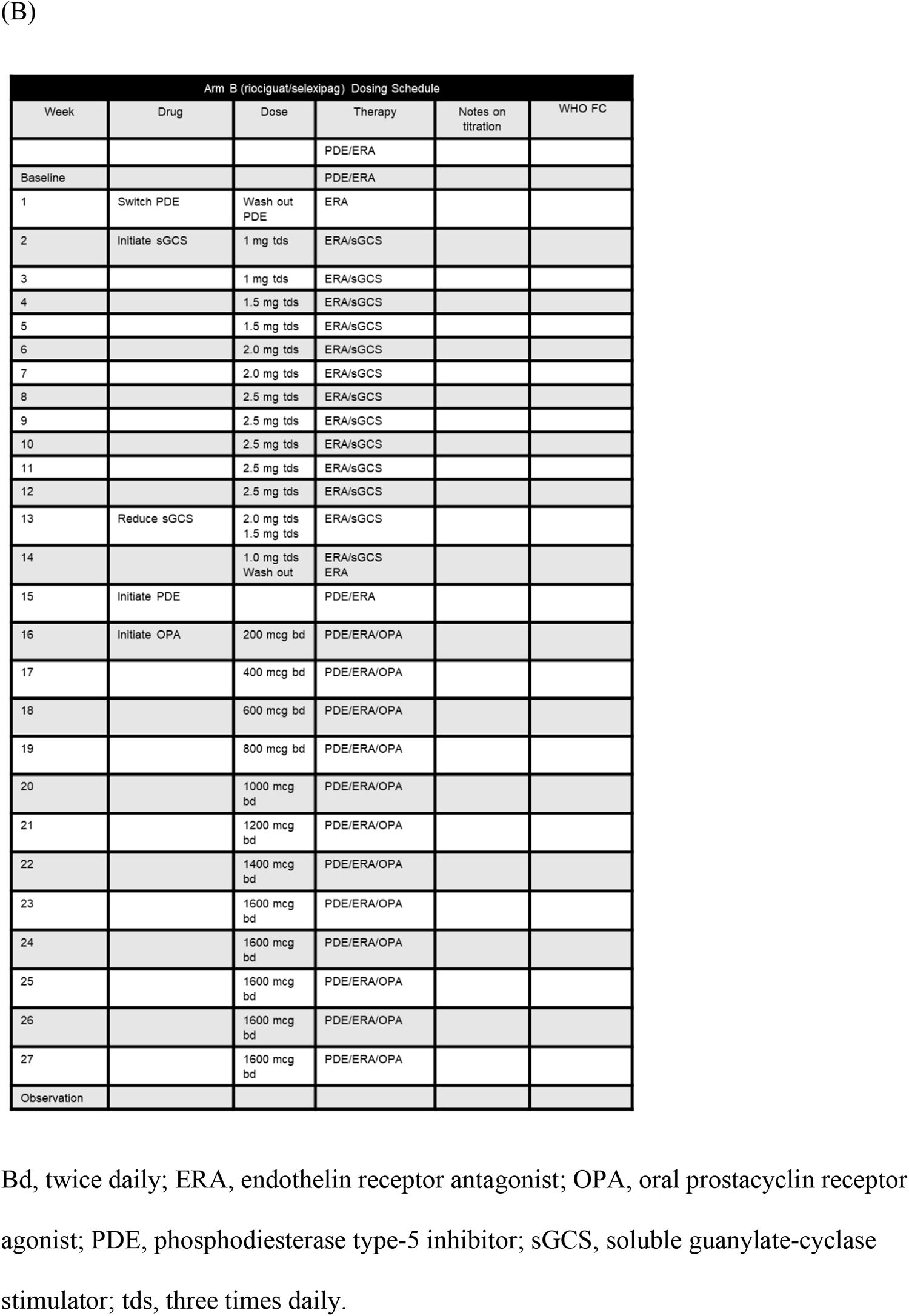
Dose escalation and de-escalation protocol for (A) Arm A and (B) Arm B.

| Arm A (selexipag/riociguat) Dosing Schedule |  |  |  |  |  |
| --- | --- | --- | --- | --- | --- |
| Week | Drug | Dose | Therapy | Notes on titration | WHO FC |
| Screening |  |  | PDE/ERA |  |  |
| Implantation |  |  | PDE/ERA |  |  |
| 1 | Initiate OPA | 200 mcg bd | PDE/ERA/OPA |  |  |
| 2 |  | 400 mcg bd | PDE/ERA/OPA |  |  |
| 3 |  | 600 mcg bd | PDE/ERA/OPA |  |  |
| 4 |  | 800 mcg bd | PDE/ERA/OPA |  |  |
| 5 |  | 1000 mcg bd | PDE/ERA/OPA |  |  |
| 6 |  | 1200 mcg bd | PDE/ERA/OPA |  |  |
| 7 |  | 1400 mcg bd | PDE/ERA/OPA |  |  |
| 8 |  | 1600 mcg bd | PDE/ERA/OPA |  |  |
| 9 |  | 1600 mcg bd | PDE/ERA/OPA |  |  |
| 10 |  | 1600 mcg bd | PDE/ERA/OPA |  |  |
| 11 |  | 1600 mcg bd | PDE/ERA/OPA |  |  |
| 12 |  | 1600 mcg bd | PDE/ERA/OPA |  |  |
| 13 | Reduce OPA | 1200 mcg bd<br>800 mcg bd<br>600 mcg bd | PDE/ERA/OPA |  |  |
| 14 |  | 400 mcg bd<br>200 mcg bd | PDE/ERA/OPA |  |  |
| 15 |  |  | PDE/ERA |  |  |
| 16 | Switch PDE | Wash out PDE | ERA |  |  |
| 17 | Initiate sGCS | 1 mg tds | ERA/sGCS |  |  |
| 18 |  | 1 mg tds | ERA/sGCS |  |  |
| 19 |  | 1.5 mg tds | ERA/sGCS |  |  |
| 20 |  | 1.5 mg tds | ERA/sGCS |  |  |
| 21 |  | 2.0 mg tds | ERA/sGCS |  |  |
| 22 |  | 2.0 mg tds | ERA/sGCS |  |  |
| 23 |  | 2.5 mg tds | ERA/sGCS |  |  |
| 24 |  | 2.5 mg tds | ERA/sGCS |  |  |
| 25 |  | 2.5 mg tds | ERA/sGCS |  |  |
| 26 |  | 2.5 mg tds | ERA/sGCS |  |  |
| 27 |  | 2.5 mg tds | ERA/sGCS |  |  |
| Observation |  |  |  |  |  |

(B)
| Arm B (riociguat/selexipag) Dosing Schedule |  |  |  |  |  |
| --- | --- | --- | --- | --- | --- |
| Week | Drug | Dose | Therapy | Notes on titration | WHO FC |
|  |  |  | PDE/ERA |  |  |
| Baseline |  |  | PDE/ERA |  |  |
| 1 | Switch PDE | Wash out PDE | ERA |  |  |
| 2 | Initiate sGCS | 1 mg tds | ERA/sGCS |  |  |
| 3 |  | 1 mg tds | ERA/sGCS |  |  |
| 4 |  | 1.5 mg tds | ERA/sGCS |  |  |
| 5 |  | 1.5 mg tds | ERA/sGCS |  |  |
| 6 |  | 2.0 mg tds | ERA/sGCS |  |  |
| 7 |  | 2.0 mg tds | ERA/sGCS |  |  |
| 8 |  | 2.5 mg tds | ERA/sGCS |  |  |
| 9 |  | 2.5 mg tds | ERA/sGCS |  |  |
| 10 |  | 2.5 mg tds | ERA/sGCS |  |  |
| 11 |  | 2.5 mg tds | ERA/sGCS |  |  |
| 12 |  | 2.5 mg tds | ERA/sGCS |  |  |
| 13 | Reduce sGCS | 2.0 mg tds<br>1.5 mg tds | ERA/sGCS |  |  |
| 14 |  | 1.0 mg tds<br>Wash out | ERA/sGCS<br>ERA |  |  |
| 15 | Initiate PDE |  | PDE/ERA |  |  |
| 16 | Initiate OPA | 200 mcg bd | PDE/ERA/OPA |  |  |
| 17 |  | 400 mcg bd | PDE/ERA/OPA |  |  |
| 18 |  | 600 mcg bd | PDE/ERA/OPA |  |  |
| 19 |  | 800 mcg bd | PDE/ERA/OPA |  |  |
| 20 |  | 1000 mcg bd | PDE/ERA/OPA |  |  |
| 21 |  | 1200 mcg bd | PDE/ERA/OPA |  |  |
| 22 |  | 1400 mcg bd | PDE/ERA/OPA |  |  |
| 23 |  | 1600 mcg bd | PDE/ERA/OPA |  |  |
| 24 |  | 1600 mcg bd | PDE/ERA/OPA |  |  |
| 25 |  | 1600 mcg bd | PDE/ERA/OPA |  |  |
| 26 |  | 1600 mcg bd | PDE/ERA/OPA |  |  |
| 27 |  | 1600 mcg bd | PDE/ERA/OPA |  |  |
| Observation |  |  |  |  |  |

Bd, twice daily; ERA, endothelin receptor antagonist; OPA, oral prostacyclin receptor agonist; PDE, phosphodiesterase type-5 inhibitor; sGCS, soluble guanylate-cyclase stimulator; tds, three times daily.

**Supplementary Figure 1.**
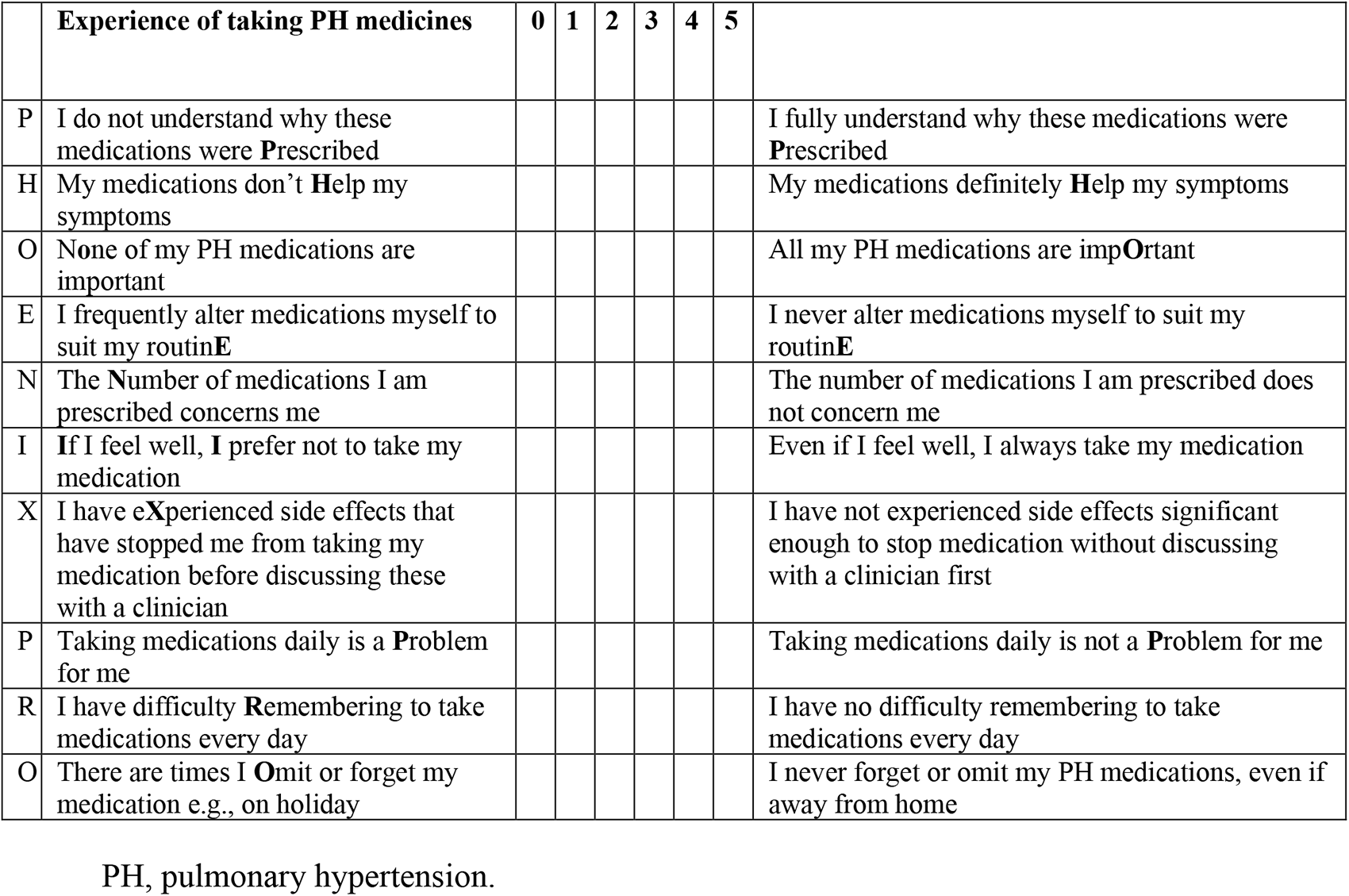
Phoenix PRO questionnaire

