## Supplementary tables and figures for "Pulmonary Hypertension: Intensification and Personalisation of Combination Rx (PHoenix): A phase IV randomised trial for the evaluation of dose-response and clinical efficacy of riociguat and selexipag using implanted technologies"

**Supplementary appendix:**

**Supplementary Table 1.** Schedule of events

Assessments at screening and baseline need only be completed once if no change in clinical condition between visits. Dosing schedule follows on next page.
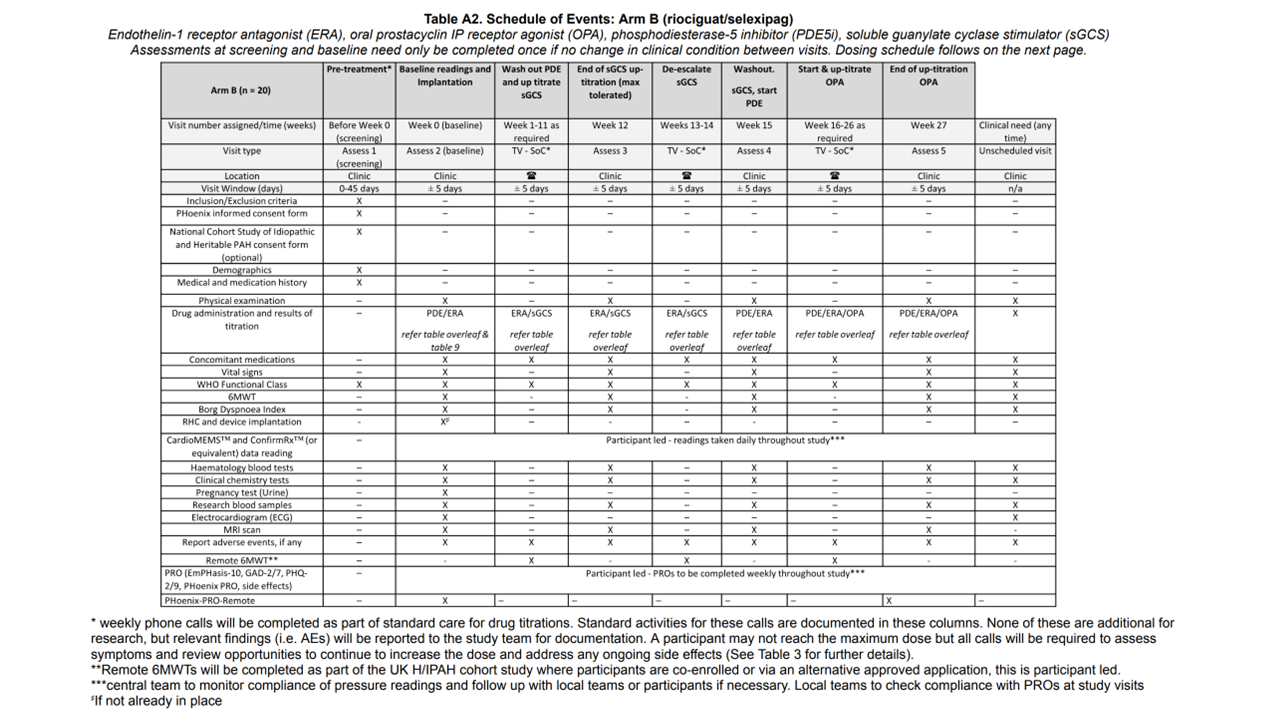

AEs, adverse events; ERA, endothelin receptor antagonist; GAD, generalised anxiety disorder questionnaire; MRI, magnetic resonance imaging; OPA, oral prostaglandin receptor agonist; PDE, phosphodiesterase type-5 inhibitor; PHQ, patient health questionnaire; PRO, patient-reported outcomes; RHC, right heart catheterisation; SoC, standard of care; sGCS, soluble guanylate-cyclase stimulator; TV, telephone visit; 6MWT, six-minute walk distance; WHO, World Health Organisation.

*Weekly phone calls will be completed as part of standard care for drug titrations. Standard activities for these calls are documented in these columns. None of these are additional for research, but relevant findings (i.e., AEs) will be reported to the study team for documentation. A participant may not reach the maximum dose, but all calls will be required to assess symptoms and review opportunities to continue to increase the dose and address and ongoing side effects.

**Remote 6MWTs will be completed as part of the UK H/IPAH cohort study where participants are co-enrolled or via an alternative approved application; this is participant led.

***Central team to monitor compliance of pressure readings and follow up with local teams or participants if necessary. Local teams to check compliance with PROs at study visits.

^$^If not already in place.

**Supplementary Table 2.** Dose escalation and de-escalation protocol for (A) Arm A and (B) Arm B

(A)

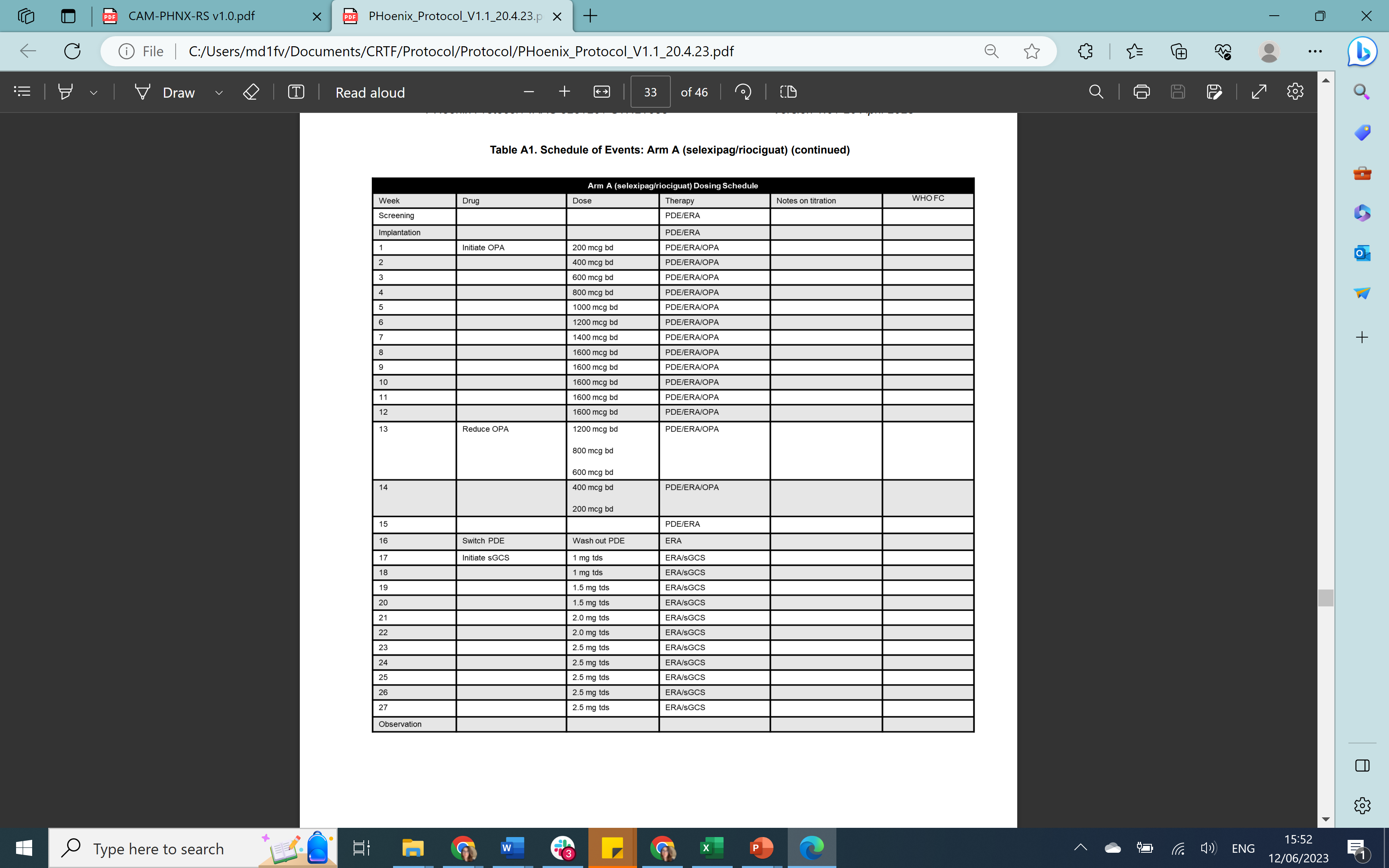

(B)

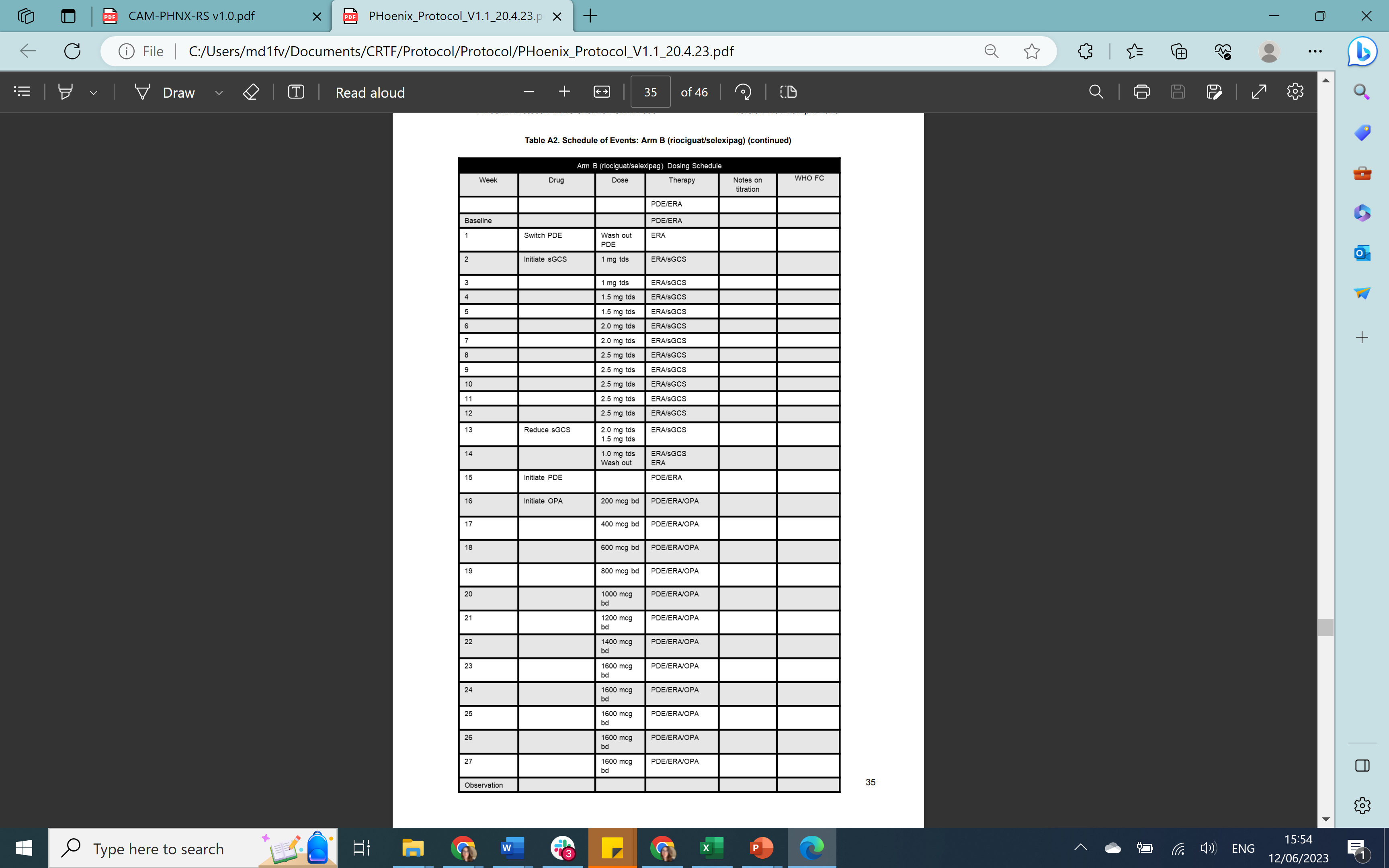

Bd, twice daily; ERA, endothelin receptor antagonist; OPA, oral prostacyclin receptor agonist; PDE, phosphodiesterase type-5 inhibitor; sGCS, soluble guanylate-cyclase stimulator; tds, three times daily.

**Supplementary Figure 1.** Phoenix PRO questionnaire (10-items), with data collected through Atom5 mobile application or via telephone.

|  | **Experience of taking PH medicines** | **0** | **1** | **2** | **3** | **4** | **5** |  |
| --- | --- | --- | --- | --- | --- | --- | --- | --- |
| P | I do not understand why these medications were **P**rescribed |  |  |  |  |  |  | I fully understand why these medications were **P**rescribed |
| H | My medications don’t **H**elp my symptoms |  |  |  |  |  |  | My medications definitely **H**elp my symptoms |
| O | N**o**ne of my PH medications are important |  |  |  |  |  |  | All my PH medications are imp**O**rtant |
| E | I frequently alter medications myself to suit my routin**E** |  |  |  |  |  |  | I never alter medications myself to suit my routin**E** |
| N | The **N**umber of medications I am prescribed concerns me |  |  |  |  |  |  | The number of medications I am prescribed does not concern me |
| I | **I**f I feel well, **I** prefer not to take my medication |  |  |  |  |  |  | Even if I feel well, I always take my medication |
| X | I have e**X**perienced side effects that have stopped me from taking my medication before discussing these with a clinician |  |  |  |  |  |  | I have not experienced side effects significant enough to stop medication without discussing with a clinician first |
| P | Taking medications daily is a **P**roblem for me |  |  |  |  |  |  | Taking medications daily is not a **P**roblem for me |
| R | I have difficulty **R**emembering to take medications every day |  |  |  |  |  |  | I have no difficulty remembering to take medications every day |
| O | There are times I **O**mit or forget my medication e.g., on holiday |  |  |  |  |  |  | I never forget or omit my PH medications, even if away from home |
